# Identifying older adults at risk for Alzheimer’s Disease based on smartphone data obtained during wayfinding in the real world

**DOI:** 10.1101/2023.06.07.23291073

**Authors:** Jonas Marquardt, Priyanka Mohan, Myra Spiliopoulou, Wenzel Glanz, Michaela Butryn, Esther Kuehn, Stefanie Schreiber, Anne Maass, Nadine Diersch

## Abstract

Alzheimer’s disease (AD), as a leading cause for disability and death in old age, represents a major burden to healthcare systems worldwide. For the development of disease-modifying interventions and treatments, the detection of cognitive changes at the earliest disease stages is crucial. Recent advancements in mobile consumer technologies provide new opportunities to collect multi-dimensional data in real-life settings to identify and monitor at-risk individuals. Based on evidence showing that deficits in spatial navigation might constitute one of the earliest AD symptoms, we assessed whether older adults who report subjective cognitive decline (SCD) but score normally on neuropsychological assessments show differences in smartphone-assisted wayfinding behavior compared with cognitively healthy older and younger adults. Guided by a mobile application, participants had to find locations along a short route on the medical campus of the Magdeburg university. We show that performance measures that were extracted from GPS and user input data distinguish between the groups. In particular, the number of orientation stops was predictive of the SCD status in older participants. Our data suggest that cognitive changes, associated with an elevated AD risk, can be inferred from smartphone data, collected during a brief episode of an everyday behavior.

## Introduction

Currently, about 32 million people around the world are affected by Alzheimer’s disease (AD) and an additional 69 (315) million are estimated to be in the prodromal (preclinical) stage of the disease (Gustavsson et al., 2022). Moreover, the prevalence of AD is expected to rise up to 153 million by 2050, due to population growth and rising life expectancies in many countries (Nichols et al., 2022). This constitutes a major burden on societies and healthcare systems by causing enormous direct (e.g., skilled nursing or professional medical care) as well as indirect (e.g., informal caregivers) costs (Nichols et al., 2019; Wong, 2020).

Until now, there are no treatments available to cure the disease (Gauthier et al., 2016), although several drugs are showing promising results in current late-stage clinical trials. For example, the anti-amyloid antibody *lecanemab* has recently been shown to slow down cognitive decline in older adults at early AD stages (mild cognitive impairment or mild dementia due to AD) compared with a placebo group (van Dyck et al., 2023). This necessitates the development of novel diagnostic tools that measure cognitive functioning in individuals who are still asymptotic in standard neuropsychological assessments but might already show subtle cognitive changes that are associated with the accumulation of amyloid and tau in certain brain areas (Hampel et al., 2022). Identifying individuals who are at-risk for AD as early as possible will further help the implementation of disease-modifying interventions to extend the time they can live independently in their community.

In recent years, as mobile and wearable consumer technologies are becoming widely accessible and are also increasingly adopted by older age groups (Faverio, 2022), the interest in digital cognitive assessments for a cost-efficient and easy-to-administer diagnosis of neurological diseases is rapidly growing (Gillan & Rutledge, 2021; Kourtis et al., 2019; Öhman et al., 2021; Piau et al., 2019). One advantage is the possibility to collect data remotely and thereby increasing the ecological validity of the test results (Öhman et al., 2021; Schlemmer & Desrichard, 2018). In addition, multi-dimensional data from different sensors of the devices can be recorded at a high frequency, which enables the detection of complex multivariate changes in behavior that evolve with disease progression (Buegler et al., 2020; Meier et al., 2021). With respect to AD, first promising attempts were made to utilize digital tools for the detection of AD-related episodic memory decline, which is linked to computations in sub-regions of the medial temporal lobe (MTL) where tau typically starts to accumulate many years before the clinical stage of AD manifests (Berron et al., 2021, 2022; Papp et al., 2017).

Another cognitive ability, which is severely compromised in AD patients and linked to neuronal resources in the MTL, is the ability to form a spatial representation of the environment, to determine one’s own location, and to navigate successfully from one place to another (i.e., spatial navigation; Coughlan et al., 2018; Lester et al., 2017; Lithfous et al., 2013; Wolbers et al., 2014). Evidence from a growing number of experimental studies, often assessing navigational performance using virtual reality (VR) setups, indicates that spatial navigation tests may show a higher sensitivity and specificity for identifying individuals at-risk for AD than episodic memory tests (Coughlan et al., 2018). For example, Bierbrauer et al. (2020) reported that the ability to estimate the current position in the environment based on information about previous positions (i.e., path integration), which is linked to computations of spatially-tuned cells (grid cells) in the entorhinal cortex, is compromised in adults who are at genetic risk for AD (APOE e4 carriers, see also Kunz et al., 2015).

In a similar vein, Chen et al. (2021) found that individuals, who report subjective cognitive decline (SCD) but do not yet show any impairments in conventional episodic memory tests, are performing worse in a virtual navigation task and show reduced functional connectivity in relevant brain areas, relative to healthy controls. Individuals with SCD have an increased risk for progression to mild cognitive impairment (MCI) and AD (Jessen, 2014; Kielb et al., 2017; Slot et al., 2019; Snitz et al., 2018) and SCD has been related to increased severity of AD pathology in the brain, especially in areas of the MTL (Amariglio et al., 2012; Buckley et al., 2017; Perrotin et al., 2012). However, using VR technology for diagnostic purposes requires training and supervision, is cost-intensive, and testing participants in VR settings poses additional challenges such as the discrepancy between visual and body-based cues, which are both important for efficient navigation. Moreover, the susceptibility to motion sickness is increased in older age groups, further limiting the diagnostic applicability of VR (Diersch et al., 2021). Moreover, most of the previous studies investigating changes in spatial navigation focused on accuracy (e.g., pointing errors) as outcome measure, without assessing the informative value of movement trajectories. First evidence from human and animal studies, however, suggests that health-related information can be inferred from movement trajectories. For example, Coughlan et al. (2019) showed that APOE e4 carriers travel longer distances in the mobile wayfinding game “SeaHeroQuest” compared with non-carriers (Coughlan et al., 2019; for similar findings in rodent models of AD see Ying et al., 2022).

Here, we introduce a novel smartphone-assisted wayfinding task to investigate the diagnostic value of movement trajectories and related information in different participant groups during navigation in the real world. It will be determined which patterns in the data might indicate subtle changes in cognition that are potentially related to an increased risk to develop AD and consequently could serve as a digital footprint for the earliest signs of the disease. More precisely, we assessed navigational abilities in healthy younger and older participants as well as in older adults who report subjective cognitive decline (SCD) during a typical everyday task, namely, finding places in the immediate surroundings, guided by our in-house developed mobile application “Explore”. In this way, we circumvent limitations of VR-based spatial navigation tasks. Moreover, in contrast to existing digital assessments of AD-related cognitive impairment, we consider passive data (e.g., GPS data) in combination with active data (e.g., user inputs) as outcome measures (cf., Fillekes et al., 2018). The potential of passive data has recently been substantiated by Ghosh et al. (2022) who equipped a group of AD patients and healthy controls with a GPS tracker and recorded their movement trajectories whenever they left their home (accompanied as well as unaccompanied) over a period of two weeks. The authors extracted multiple parameters from the GPS data (e.g., distance from home, entropy, duration of stops, segment similarity and complexity) and showed that some of these parameters differed between the two groups and could be used to predict the participant’s cognitive health status. Whether this is similarly the case in individuals who are at risk for AD but are still clinically normal remains unknown.

Based on findings that navigating to familiar destinations remains relatively unimpaired with advancing age (Merriman et al., 2016; Rosenbaum et al., 2012), we asked our participants to find locations, which were not known to them prior to testing and which were unlikely to be part of their everyday navigation routes, on the relatively confined medical campus area of the Magdeburg university. In line with previous studies, differences between the three groups in our study might become particularly evident for the covered distances during wayfinding (cf. Coughlan et al., 2019). Alternatively, group difference might emerge on measures that capture the participants’ cognitive processing demands and uncertainty about where to go in order to reach a destination, in line with findings showing that impairments in spatial navigation may lead to behaviors aiming at lowering cognitive efforts during task performance (i.e., viewing the map more often during wayfinding or briefly stopping during navigation in order to orient; cf., Taillade et al., 2013). Answering these questions will offer critical novel information on how to use mobile data to detect behavioral patterns that are potentially associated with AD-related cognitive decline.

## Results

In this study, findings are reported from 24 younger adults, 25 cognitively healthy older adults, and 23 older adults who reported subjective cognitive decline (SCD) but performed normal in the consortium to establish a registry for Alzheimer’s disease test battery (CERAD; Welsh et al., 1994). The CERAD is an established neuropsychological assessment to detect AD-related cognitive decline, consisting of several subtests that measure different aspects of cognitive functioning (e.g., verbal fluency, episodic memory, executive functioning). Participants performed a mobile wayfinding task on the campus area around the German Center for Neurodegenerative Diseases (DZNE) in Magdeburg, Germany. Guided by our newly developed smartphone application “Explore”, participants were asked to walk from the DZNE to five salient buildings on the campus (points-of-interest, Figure 1a), while their GPS data were recorded (latitude, longitude, and timestamp). The points-of-interest (POI) had to be found consecutively, and the whole route covered approximately 820 m. At the beginning of each walking track, participants saw a map on the smartphone, showing their own location and the POI location as well as a picture of the POI (Figure 1b). Participants were instructed to close the map before they started walking and to find the POI independently (Figure 1c). When the participant walked more than 8 meters or after 18 seconds without interaction on screen, the map closed automatically. Participants had the possibility to view the map again, if needed, and the number of times they called this help function was recorded for each track (Figure 1d). A QR code, which was placed at the entrance door of the target POI, had to be scanned with the phone camera once the participant arrived at the POI. This confirmed the completion of the current track and initiated the corresponding procedure for the next track (Figure 1e). At the beginning, participants received a short practice of the task to get familiar with the app and the whole task was completed once participants returned to the DZNE after finding the five POIs. All of the participants were smartphone owners, did not report any mobility impairments and possessed comparable levels of familiarity with the campus area, as assessed in a screening session before testing (Table 1).

**Figure 1.**
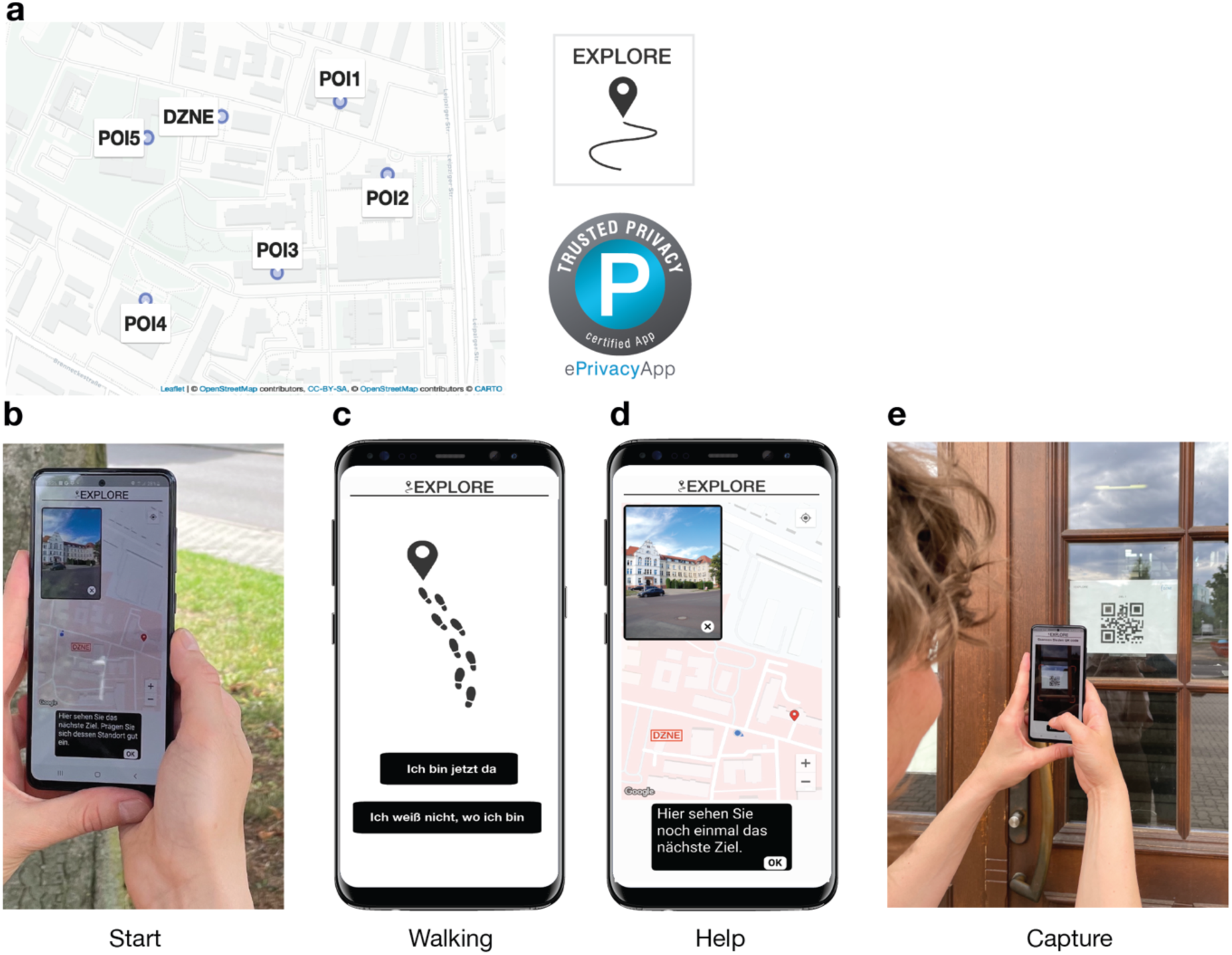
Mobile wayfinding task implemented in the “Explore” app. (a) Starting from the DZNE, participants had to find five points-of-interest (POIs) on the surrounding campus area. (b) For each POI, a map was displayed at the start of the track, showing the current location of the participant and the POI location as well as a picture of the POI (start phase). (c) During walking, the map was not shown (walking phase). (d) If needed, participants could view the map again by calling the help function (help phase). (e) A track was successfully completed when a QR code, which was placed at the entrance door of the POI, was scanned with the phone camera (capture phase).

**Table 1.** Sample characteristics (descriptives; mean scores ± SD) of the three participant groups (YA: younger adults; OA: healthy older adults; SCD: older adults with subjective cognitive decline). The CERAD composite score was calculated using the age-, gender-, and education-corrected z-scores from six different sub-tests (Boston Naming Test, verbal fluency, word list learning, word list recall, word list savings, and constructional praxis, Chandler et al., 2005). The groups did not differ in the listed attributes, all p ≥ .112 (age differences between healthy older adults and older adults with SCD were tested using a Welch two-sample t-test; gender differences using a χ2 test; differences in campus familiarity using a Kruskal-Wallis rank-sum test; Life-space assessment score differences using an analysis of variance).

|  | YA | OA | SCD |
| --- | --- | --- | --- |
| n | 24 | 25 | 23 |
| Age | 24.5 ± 2.36 | 65.7 ± 4.02 | 66.2 ± 6.54 |
| No of female | 12 | 13 | 12 |
| Campus familiarity<br>(max. score: 28) | 12.7 ± 9.15 | 14.1 ± 8.77 | 8.9 ± 6.27 |
| Life-Space Assessment<br>(max. score: 120) | 81.0 ± 12.1 | 84.2 ± 16.0 | 84.5 ± 14.1 |
| Cognitive screening<br>scores | -- | MoCA: 28.5 ± 1.12 | MMSE: 28.8 ± 1.31<br>CERAD: 0.17 ± 0.54<br>min = -0.82 |

### Differences in GPS trajectories do not distinguish between participant groups

To determine the potential of the smartphone data to distinguish between the three participant groups, we first quantified relative distances between individual GPS trajectories by using dynamic time warping (DTW; Cleasby et al., 2019; Senin, 2009), while focusing on the trajectories during the walking phases. The resulting DTW dissimilarity matrix was submitted into a k-medoids clustering analysis to identify subgroups of participants who exhibited similar wayfinding styles, and it was tested how well the obtained clusters represent the three participant classes. This analysis showed that our sample is best described by three wayfinding clusters. Most members of the first cluster (n = 35; average distance to the cluster medoid = 0.08) were walking directly from POI to POI or showed only minor deviations from the most direct path to the POIs (e.g., detours). Members of the second cluster (n = 24; average distance to the cluster medoid = 0.09) typically took a less direct way to the POIs, including wrong turns at some intersections. The third cluster consisted of two members who took largely different paths relative to the rest of the sample (average distance to the cluster medoid = 0.17). A prototypical trajectory for each cluster can be found in Figure 2a-c and the movement trajectories of the whole sample are provided in Supplementary Figure 1.

**Figure 2.**
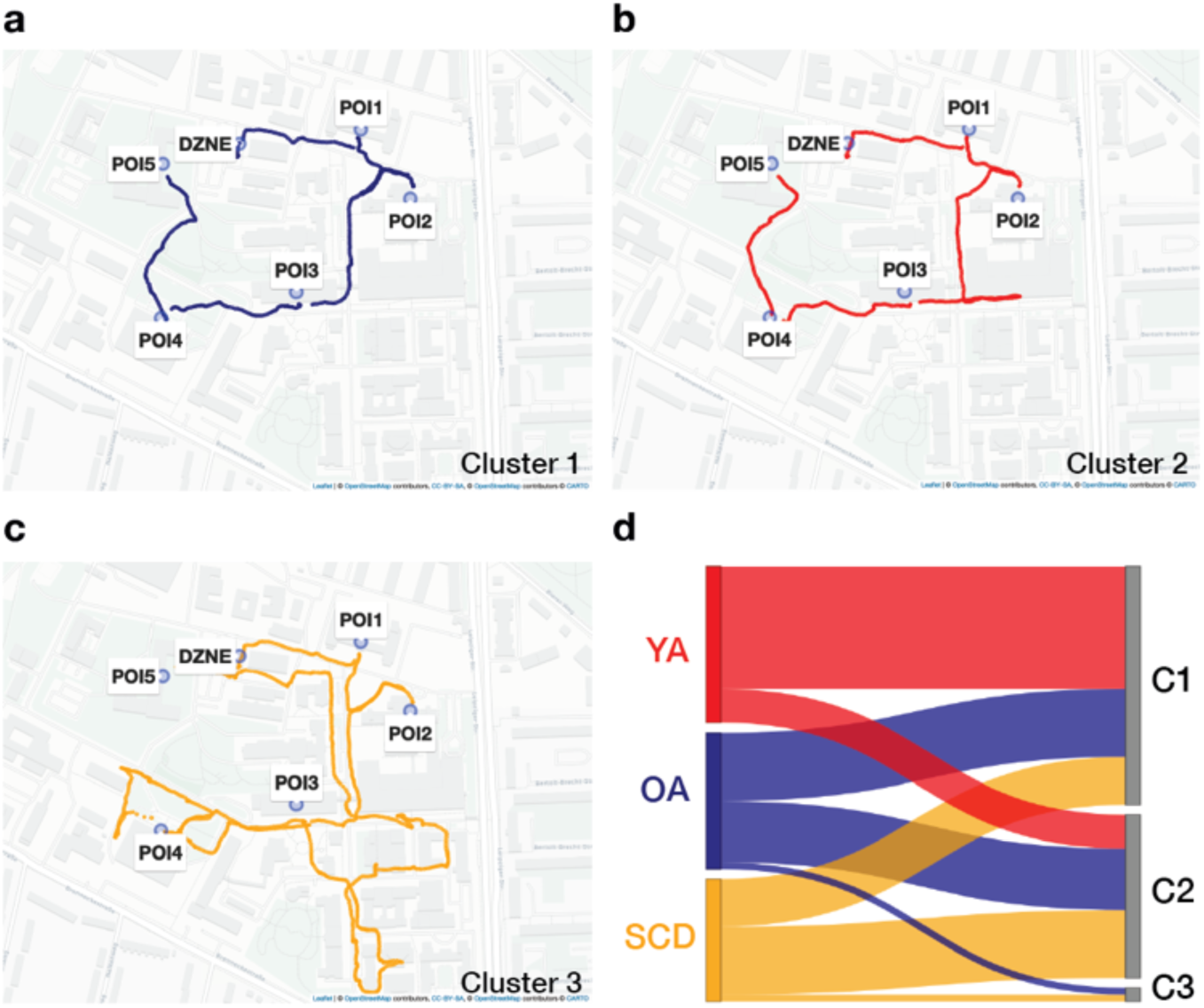
Three wayfinding styles in our sample as identified by a k-medoids clustering analysis using differences in GPS trajectories as input features. GPS trajectory of a representative member (cluster medoid) of the (a) first, (b) second, and (c) third cluster. (d) Sankey diagram showing the distribution of our participant classes in each wayfinding cluster. Red: younger adults; blue: healthy older adults; yellow: older adults with subjective cognitive decline.

The correspondence between the obtained wayfinding clusters and the three participant classes was rather low (cluster purity = 0.475, Figure 2d). The majority of younger adults (n = 18) were members of the first cluster, which also included 10 healthy older adults and seven older adults with SCD. The second cluster mainly consisted of older adults (nine healthy older adults and 10 older adults with SCD), but also included five younger adults. The third cluster consisted of one healthy older adult and one older adult with SCD. Thus, interindividual differences in the movement trajectories were not sufficient to extract age– and health-related information about the sample within the context of our task, presumably due to a high context-dependency of the data (i.e., the unique characteristics of each track).

### Group separation improves considerably when being based on aggregated performance measures

As a next step, we calculated five performance measures that were derived from the GPS data and user input data: 1) wayfinding distance; 2) wayfinding duration; 3) movement speed; 4) the number of help function calls during walking (map views); 5) the number of times the participants briefly stopped during walking (orientation stops) as a measure for increased cognitive processing demands when participants faced wayfinding difficulties (May et al., 1995; Taillade et al., 2013). A latent profile analysis (LPA; Berlin et al., 2014) was applied with these performance measures as input features to identify wayfinding performance profiles. We again tested how well the resulting profiles corresponded to the participant classes. This analysis showed that a model with three wayfinding performance profiles represented our sample best. The inspection of the single profiles with respect to their performance characteristics (Figure 3a) showed that participants expressing the first profile (n = 25) can be described as high-level performers who covered less distance, needed less time, and generally moved quicker during wayfinding. They also looked at the map less often during walking and had fewer orientation stops. The second profile was characterized by a mid-level performance on all measures (n = 29). Participants expressing the third profile (n = 7) covered more distance during walking, needed more time to complete the tracks, and moved slower. The biggest difference was evident for the number of times they called the help function and the number of the times they briefly stopped during walking, with both measures being considerably higher than in the other two profiles. Thus, participants with this profile can be described as low-level performers who had the biggest difficulties in finding the POIs.

**Figure 3.**
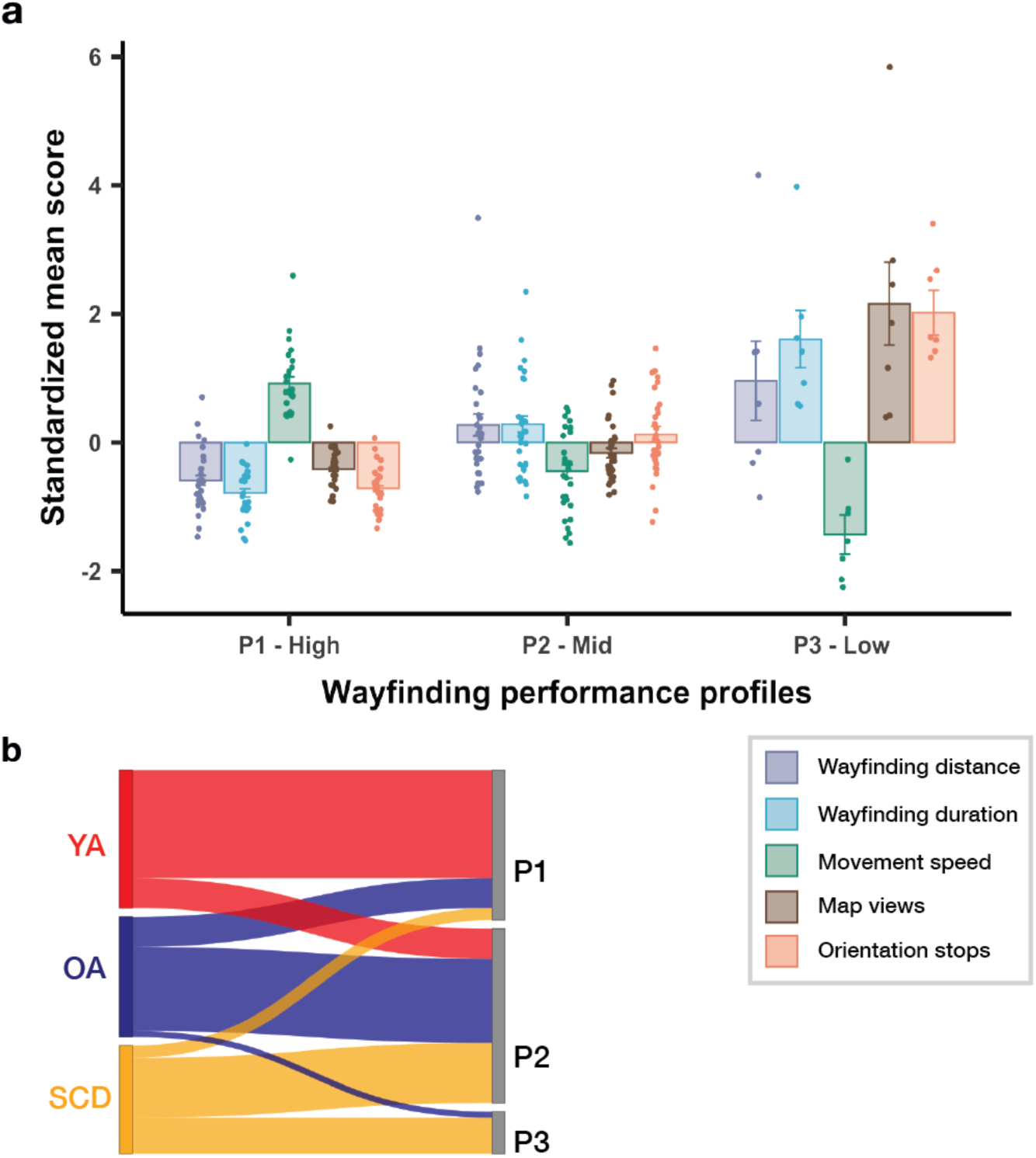
Three wayfinding performance profiles in our sample (P1: high-level; P2: mid-level; P3: low– level) as identified by a latent profile analysis (LPA) using five performance measures as input features. (a) Means (± standard error) of the z-scored performance measures for each profile (purple: wayfinding distance; light blue: wayfinding duration; green: movement speed; brown: number of map views during walking; orange: number of orientation stops). (b) Sankey diagram showing the distribution of our participant classes in each wayfinding performance profile. Red: younger adults; blue: healthy older adults; yellow: older adults with subjective cognitive decline.

Importantly, the participant classes were better represented by these wayfinding profiles than by the wayfinding clusters that we obtained from differences in the GPS trajectories (profile purity = 0.623, Figure 3b). The majority of the high-level performers were younger adults (n = 18). Only five healthy older adults and two older adults with SCD showed a similar high-performance pattern. Most of the healthy older adults (n = 14) and those with SCD (n = 10) were navigators who were characterized by a mid-level performance pattern in the context of our task. Five younger adults also fell in this category. A low-level performance was expressed by six older adults with SCD and one healthy older adult. This shows that different wayfinding performance parameters, which were extracted from GPS and user input data, are better suitable to identify age groups and older adults with SCD than differences between movement trajectories.

### The number of orientation stops differs between healthy older adults and older adults with SCD

To investigate group differences on each performance measure in more detail, we fitted linear and generalized mixed effect (LME/GME) models with gender and campus familiarity as covariates and random intercepts for participant and track to the data. This analysis showed that younger adults differed significantly from the two older participant groups on all performance measures (all p ≤ .018, see Supplementary Tables 1-5 for the detailed results of each LME and GME model). Compared with their older counterparts, they covered less distance, needed less time, had a higher movement speed, used the help function less often, and had fewer orientation stops during wayfinding (Figure 4). With respect to differences between healthy older adults and older adults with SCD, we found that older adults with SCD showed a significantly higher number of orientation stops than healthy older adults, β = 0.67, p = .007 (Figure 4e). They additionally tended to look at the map more often during walking, β = 0.82, p = .059 (Figure 4d), and tended to need more time to complete the task, β = 0.13, p = .088 (Figure 4b). This cannot be explained by walking more slowly since the average movement speed did not differ between the two older groups, β = –0.05, p = .371 (Figure 4c). Older adults with SCD also did not cover more distance than the healthy older adults, β = 0.07, p = .208 (Figure 4a).

**Figure 4.**
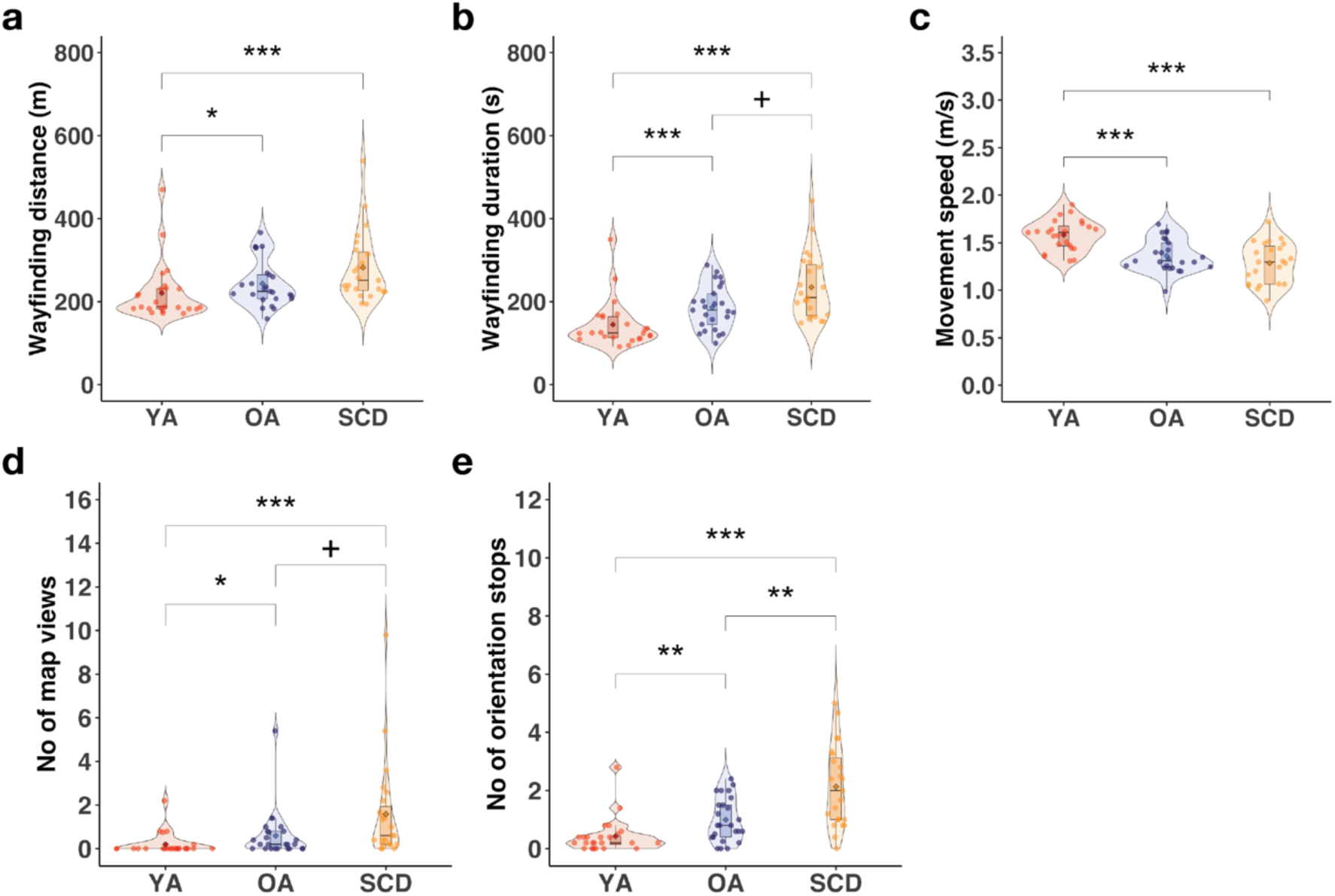
Results of the mixed effect model analyses. Performance of the three groups (red: younger adults; blue: healthy older adults; yellow: older adults with subjective cognitive decline) for the (a) wayfinding distance; (b) wayfinding duration; (c) movement speed; (d) number of map views during walking; (e) number of orientation stops (divided by the number of completed tracks). The boxplot denotes the lower and upper quartile of the measure; center line the median; whiskers the 1.5x interquartile range; dots the individual data points; diamond shape the mean; ^+^ p < .10; * p < .05; ** p < .01; *** p < .001. The detailed results of all models are listed in the Supplementary Tables 1-5.

Taken together, we found that briefly stopping in order to orient themselves differed between healthy older adults and older adults with SCD. In contrast, general walking patterns, such as the overall distance that was covered and the movement speed, were largely similar among older adults, with the former presumably also being more influenced by the specific characteristics of the tracks. This is further supported by the variability between participants and tracks that we modeled as random effects in our models, showing that differences between tracks explained more variance than individual differences for wayfinding distance, whereas the reverse was true for the number of map views and orientation stops (Supplementary Tables 1-5, see also Supplementary Figure 2 for the number of orientation stops on each track in healthy older adults and older adults with SCD).

### The number of orientation stops predicts SCD status in older adults

As a next step, we were interested whether the average number of orientation stops that differed between healthy older adults and older adults with SCD, could be used to predict SCD in older adults (n = 48). The number of orientation stops was consequently fed into a logistic regression model with SCD status as outcome variable. The logistic regression model was significant, χ^2^ (1) = 8.1, p = .004 (see Figure 5a). A higher number of orientation stops was associated with significantly higher odds of being an older adult with SCD than a healthy older adult, Odds Ratio = 2.70, 95% CI = 1.47 – 5.86, z = 2.85, p = .004. A leave-one-out (LOO) cross-validation confirmed that SCD status of unknown participants can be predicted above chance when using the predictions of the logistic regression model (accuracy = 0.67, CI = 0.53 – 0.80, chance level 0.50). Overall, 32 out of 48 individuals were correctly classified.

**Figure 5.**
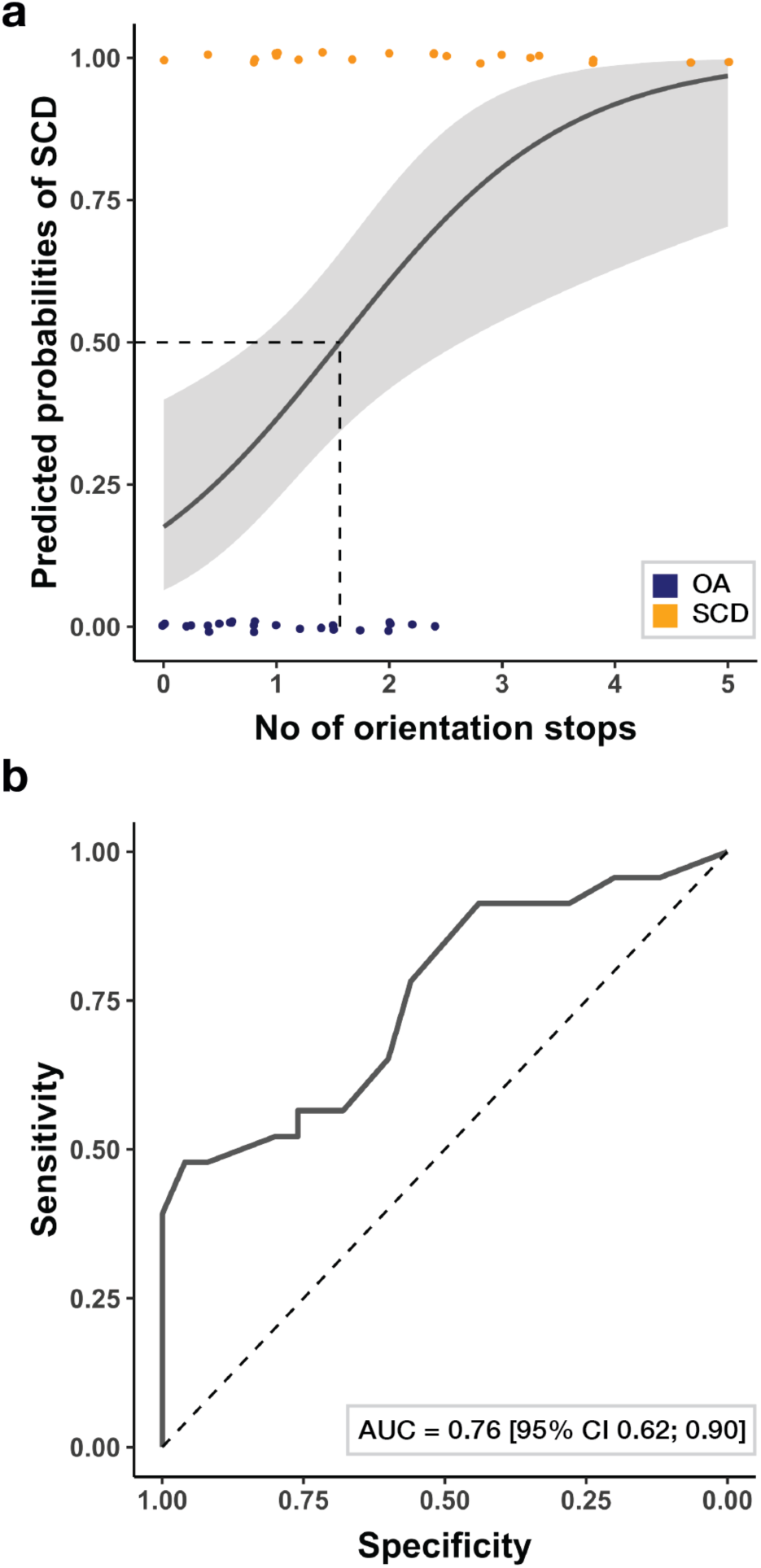
The number of orientation stops as predictor of SCD status in older adults. (a) Predicted probabilities of being classified as an older adult with SCD as estimated in a logistic regression model with the number of orientation stops as input feature. Shaded areas denote the 95% confidence intervals of the predictions and dots the individual data points (blue: healthy older adults; yellow: older adults with subjective cognitive decline). (b) Receiver Operating Characteristic (ROC) curve showing the diagnostic accuracy of the number of orientation stops for the detection of SCD in older adults.

A receiver operating characteristic (ROC) analysis showed that the number of orientation stops differentiated healthy older adults and older adults with SCD with an area under curve (AUC) of 0.757 (SE = 0.07, 95% CI: 0.62 – 0.90, Figure 5b). A maximum sensitivity of 78% could be achieved with a specificity of at least 50%, and a maximum specificity of 80% could be achieved with a sensitivity of at least 50%. Thus, our results provide evidence that the number of orientation stops during wayfinding is predictive of SCD status in older adults.

### The number of orientation stops might be linked to executive functioning in older adults with SCD

The individual number of orientation stops varied widely within older adults with SCD across tracks. To provide more insights into the cognitive processes that might be associated with this digital performance measure, we calculated in a post-hoc analysis the correlations between the number of orientation stops and the CERAD subtest scores that were available for this group (Welsh et al., 1994). This analysis showed that the number of orientation stops was moderately correlated with the MMSE score as a measure for general cognitive functioning (r = –0.33), as well as with the Constructional Praxis Recall (r = –.32) and Savings test scores (r = –.031), which assess visual memory functioning. The highest correlation was obtained for the Trail Making A/B score as a measure for executive functioning (task switching, Arbuthnott & Frank, 2000) suggesting that a better performance in this test might be linked to fewer orientation stops during wayfinding in the real world in older adults with SCD (r = –.39, Figure 6). However, none of the correlations reached statistical significance and should therefore be interpreted with caution (all p ≥ .068, see Supplementary Table 6 for all results).

**Figure 6.**
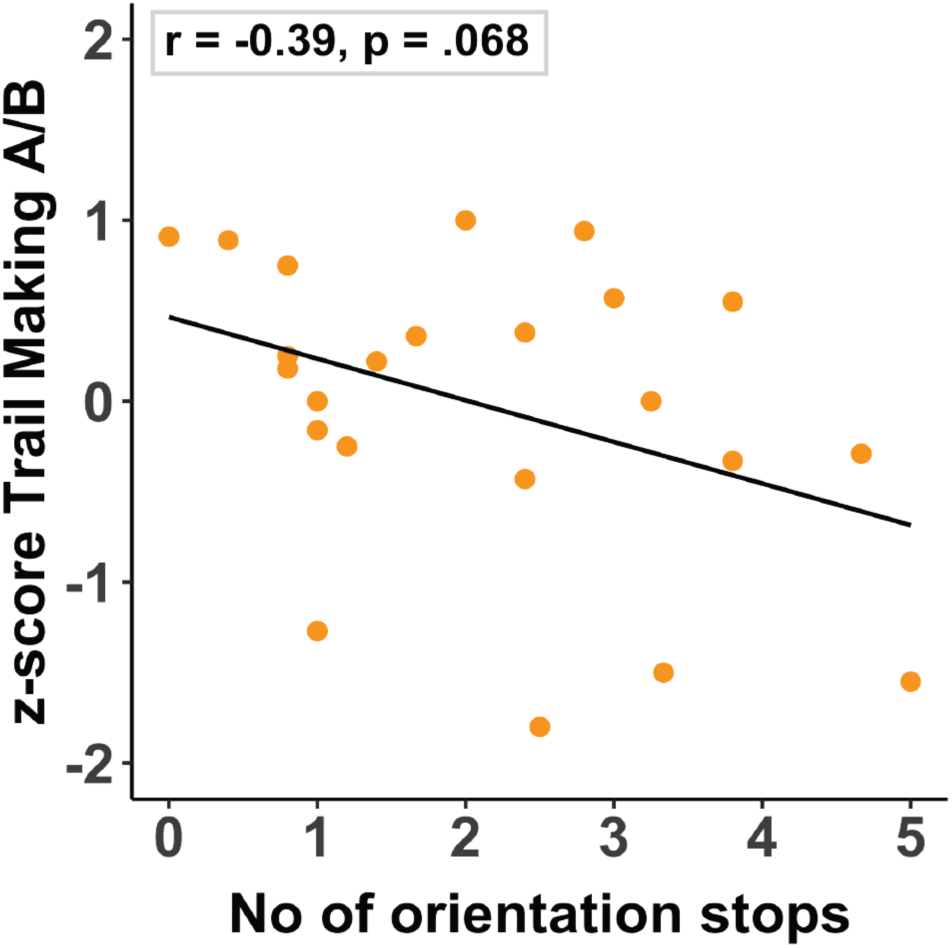
Correlation between the average number of orientation stops and the z-scored Trail Making A/B ratio of the CERAD test battery in older adults with SCD.

## Discussion

In this study, we evaluated the potential of smartphone data, recorded during wayfinding in the real world, to distinguish between healthy older adults and older adults with SCD who are known to possess a higher risk for developing AD. We show that behavioral wayfinding performance indicators, derived from GPS and user input data, contain information about the participant’s age group and SCD status. Specifically, the number of brief stops while navigating to different locations in the environment differed between groups. This effect was strong enough to predict SCD status in older participants, rendering this performance measure as a promising digital footprint for AD-related cognitive decline in real world settings. By showing that older adults with SCD can be distinguished from cognitively healthy older adults during a mobile wayfinding task that was completed in less than half an hour, we extend on findings by Ghosh et al. (2022) who reported that GPS data differ between AD patients and healthy controls when tracking their movement trajectories over a period of two weeks. Moreover, the number of orientation stops identified older adults with SCD with an AUC of 0.76, which is comparable to assessments of navigational performance in virtual environments (Chen et al., 2021; Coughlan et al., 2019). However, in contrast to these studies, we identified these individuals on the basis of digital data obtained during a short and remotely performed everyday task, which poses a major advantage for a potential future application in primary healthcare.

In contrast to previous VR navigation studies, we did not observe differences between healthy older adults and older adults at-risk for AD with respect to the distances that were covered during wayfinding (Bierbrauer et al., 2020; Chen et al., 2021; Coughlan et al., 2019). This might be related to the characteristics of the virtual environments in these studies, which often entailed relatively unconstrained spaces (e.g., virtual arenas) where participants have to rely predominantly on self-motion cues in order to estimate their position in the environment. This process (path integration) has been linked to computations of grid cells in the entorhinal cortex, which are affected by AD pathology at an early stage (Braak et al., 2006; Kunz et al., 2015; Segen et al., 2022). In the real world, in contrast, participants can additionally draw on various visual cues (e.g., landmarks) to aid their performance (cf., Huffman & Ekstrom, 2021) and their movement paths are naturally constrained by streets and barriers such as vegetation and objects (e.g., cars or fences), which might obscure potential deficits in path integration. This shows that results from laboratory-based studies using VR cannot be transferred on par to real-world settings and that cognitive change might manifest differently in real-world behavior. This is further supported by the finding that differences in the movement trajectories were not sufficient in order to clearly separate the groups, presumably due to the specific characteristics of the single tracks that influenced the paths that were taken.

Group differences emerged instead in the number of orientation stops during wayfinding, in line with Taillade et al. (2013) who observed a higher number of brief stops in healthy older adults compared with younger adults when they were asked to reproduce a path in the real world that was previously learned in VR. The authors concluded that more orientation stops during navigation might indicate impairments in executive functioning. Intact executive functioning is an important component of efficient navigation, for example, when switching between different navigation strategies or planning a route (Colombo et al., 2017). In addition to planning a route to reach a certain destination, monitoring the progress when walking towards the destination as well as decision making processes at intersections are all processes that strongly depend on executive functioning (Moffat, 2009; Wolbers & Hegarty, 2010). Moreover, due to age-related declines in sensorimotor processing that are partly compensated by cognitive strategies in order to reduce the risk of falling during walking, fewer cognitive resources are available for other operations with advancing age in such situations (Li et al., 2018; Seidler et al., 2010). Thus, an increase in dual-task demands in a situation where the demands on cognitive processing are increased, for example, when translating information gathered from a map into concrete actions in an area that is not visited on a regular basis, might be one explanation for the higher number of orientation stops in the older age groups. This is in line with previous findings showing a detrimental effect of unsupported, active walking while solving a spatial navigation task in older compared with younger adults (Lövdén et al., 2005).

Recently, however, it has been shown that cognitive performance also seems to be maintained or even improved in some older adults in a dual-task walking condition (Patelaki et al., 2023). This suggests that the ability to reallocate neural resources in dual-task situations might be a characteristic of successful cognitive aging. In line with this, Åhman et al. (2020) observed worse performance in older adults with SCD compared with healthy older adults when they performed a dual-task that involved naming animals or months of the year backwards while walking. Similarly, Montero-Odasso et al. (2017) demonstrated that a lower gait velocity while performing a cognitive task is predictive of AD in patients with MCI. These findings suggest that AD-related wayfinding deficits in real-world settings are likely to be linked to declines in executive functioning and impairments in the processing of dual-task demands. While the older adults with SCD in the current study showed a similar movement speed as the healthy older adults and possessed comparable activity levels in their daily lives, our correlational analysis similarly indicates that the measure that differentiated between healthy older adults and those with SCD might be associated with executive functioning. The precise contribution of different cognitive processes on digital wayfinding performance parameters will be an important topic for future research.

The LPA in which five performance measures were used as input features showed that our sample was described best by three different wayfinding profiles. In terms of the overlap with our participant classes, we found that each wayfinding profile was expressed predominantly by one of the three groups, for example, most high-performing navigators were younger adults, whereas most of the low-performing navigators were older adults with SCD. However, in line with many previous studies on age-related cognitive decline, we also observed a certain amount of within-group variability with quite a few older adults with SCD showing a mid-level performance pattern, and some even being classified as high-level performers (cf., Lindenberger, 2014; Mungas et al., 2010; Nyberg & Pudas, 2019). Although the likelihood of progression to AD is increased for older adults with SCD and even stronger for older adults with SCD who are patients in memory clinics, as in our case (Slot et al., 2019), additional indicators need to be considered in order to predict who will progress to AD or not (Jessen et al., 2020). Our sample size was not large enough to assess the diagnostic accuracy of the wayfinding performance parameters in sub-groups that were stratified with respect to additional predictors of AD, which is one of the limitations of the study.

To better explain within-group heterogeneity in wayfinding performance, future studies should assess APOE e4 status, as one of the strongest predictors for the future development of AD (Corder et al., 1993; Farrer et al., 1997). Previous research showed that young APOE e4 carriers show higher wayfinding distances in virtual spatial navigation tasks (e.g., Coughlan et al., 2019) and APOE e4 status has also been linked to an increased subjective perception of cognitive decline (Zhang et al., 2017). Thus, one might hypothesize that older adults with SCD might differ from healthy older adults on additional wayfinding performance measures, depending on their genetic risk for AD. Information on biomarker status (tau and amyloid beta protein accumulation) will further help to improve the characterization of AD risk in older adults (Cullen et al., 2021; Janelidze et al., 2020), which in turn influences their navigation behavior (Coughlan et al., 2018; Lester et al., 2017). Furthermore, future studies should assess lifestyle factors such as physical fitness (Maass et al., 2015) and cognitive (Stern, 2009) as well as vascular reserve factors (Perosa et al., 2020; Vockert et al., 2021) that contribute to preserved MTL function in old age, even in the face of emerging pathology.

Taken together, we show that digital markers, extracted from smartphone data acquired during a remotely performed wayfinding task that took less than half an hour to complete, are suggestive of the cognitive health status in older adults. The results of our study are a starting point for determining how smartphone data, acquired during a frequently performed everyday behavior, can be used for the assessment of cognitive impairment in the context of AD. We circumvented common limitations of VR technology, such as the increased susceptibility to cybersickness in older adults (Diersch & Wolbers, 2019) and show that AD-related changes in wayfinding behavior in the real world might manifest differently than during wayfinding in VR. We further identified features from the data that differed between the groups and were less dependent on the characteristics of the environment (e.g., the specific track that was traveled), providing first indications how such data could be acquired independent of location. As a next step, the potential of additional features, for example, from sensor data that track fine-motor movements (del Pozo Cruz et al., 2022) or vocal information from the microphone (König et al., 2015), should be determined for a better classification of participant groups who are at risk for AD. Multimodal data from smartphones and wearables have the potential to better account for different developmental trajectories and distinct subtypes that characterize the disease (Ferreira et al., 2020; Vogel et al., 2021).

With the rising adoption of smartphones and wearables in older age groups (Faverio, 2022), data from mobile applications like the “Explore” app could ultimately be used as a screening tool to stratify subjects with regard to the need of extended cognitive and clinical diagnostics at all and to which detailedness. Using digital wayfinding performance measures might support the identification of individuals who may benefit from pharmacological treatments (van Dyck et al., 2023) or behavioral interventions (Livingston et al., 2020; Ngandu et al., 2015). Based on evidence showing that spatial navigation training enhances cognition and maintains MTL function in older adults (Lövdén et al., 2012; McLaren-Gradinaru et al., 2020; Montana et al., 2019), mobile tools like the “Explore” app might further provide the means to combine the positive effects of lifestyle interventions, for example, physical exercise (Maass et al., 2015) and cognitive training (Ball et al., 2002; Schmiedek et al., 2010; Uttal et al., 2013) to slow down the progression of AD-related cognitive decline.

## Material and Methods

### Sample

In total, 72 participants took part in the study (24 younger adults, 25 cognitively healthy older adults, and 23 older adults with SCD; see Table 1 for the characteristics of the sample). Older adults with SCD were transferred to the study from the DZNE memory clinic and the definition of SCD followed the criteria as proposed by Jessen et al. (2014): 1) self-report of lowered cognitive capacity in contrast to a formerly normal capacity that is not caused by a severe event; 2) unimpaired performance within the age-, gender-, and education-matched norms on neuropsychological tests used to assess MCI or prodromal AD; 3) exclusion of MCI, prodromal AD, or any other type of dementia; 4) the self-perceived decline of cognition cannot be attributed to a psychiatric, neurological, or medical disease apart from AD nor to medication or substance abuse. Their cognitive health status was assessed by the consortium to establish a registry for Alzheimer’s disease test battery (CERAD; Welsh et al., 1994), including the Mini-Mental State Examination (MMSE; Folstein et al., 1975). The remaining participants were recruited from the DZNE participant database. To be included as a cognitively healthy older adult, a score higher than 23 had to be obtained in the Montreal Cognitive Assessment (MoCA; Luis et al., 2009; Nasreddine et al., 2005). All participants had normal or corrected-to-normal vision and none of them reported a history of psychiatric, neurological, or motoric diseases or use of medication that might affect task performance. All participants were community-dwelling individuals with no major mobility impairments as determined by the German version of the Life-Space-Assessment (LSA; Baker et al., 2003; Mümken et al., 2021). This questionnaire measures the frequency of activities in five different life-spaces within the past month, ranging from the participant’s home to places outside of town, and further considers the level of independence with which these activities were completed. In addition, all participants were smartphone owners and, thus, can be considered as being experienced in using mobile devices. Participants provided informed consent and were paid for their participation in accordance with the local ethics committee.

### Assessment of the prior knowledge of the environment

Familiarity with the campus area around the DZNE Magdeburg was assessed prior to testing by using a self-report questionnaire that assessed the number of previous visits on the campus, their frequency and the number of buildings that were usually visited. The questionnaire further contained a short spatial memory test about the campus area, consisting of three subtests (Supplementary Figure 3). In the first test, participants had to indicate from a list of pictures showing 12 campus buildings (including the five POIs of the mobile wayfinding task), which of the buildings they recognize (maximum score: 12). In the second test, four picture triplets of the 12 buildings were presented. Here, participants had to choose for each triplet, which of the two lower buildings lies closer to the upper reference building (maximum score: 4). They also had the option to indicate that they don’t know the answer. In the third test, they saw a map of the campus with red dots at different locations and had to assign the 12 buildings to the corresponding dots, in this way identifying their location (maximum score: 12). The items from the spatial memory test showed an acceptable internal consistency (Cronbach’s α = 0.75) and the sum of correct answers served as total familiarity score in the analyses. Overall, our sample showed an intermediate familiarity with the environment and, importantly, there were no significant differences in the total familiarity score between the three groups, χ^2^(2) = 4.38, p = .112 (Table 1).

### Mobile wayfinding task

The mobile wayfinding task was implemented within the smartphone application “Explore” that was conceptualized and designed by Nadine Diersch and realized by Intenta GmbH (https://www.intenta.de/en). For testing, the app was installed on two identical, DZNE-owned phones (Samsung A51) to ensure that all participants performed the task under the same conditions (e.g., in terms of display size or performance of the phone) and were not distracted by incoming calls or messages. The app was certified with the “ePrivacyApp” seal by ePrivacy (https://www.eprivacy.eu/en/privacy-seals/eprivacyapp/) to meet standard European data protection and security requirements.

In the mobile wayfinding task, five distinct buildings (points-of-interest, POI) had to be found on the campus area around the DZNE, with the DZNE serving as start and end point. This resulted in six different walking tracks that could be completed along a route that covered approximately 820 m. At the beginning of each track, a map was displayed that showed the location of the target POI (start phase). In addition, the participant’s location was displayed dynamically as a blue dot with a small arrow, indicating the pointing direction of the phone. For all tracks, the DZNE was marked on the map for reference. The visible section of the map was freely movable by the participant via dragging and the map could be zoomed via pinching. In the upper left corner of the display, a picture of the target POI was shown that could be closed and reopened. The start phase ended automatically, when the participant walked more than 8 m or after a time-out of 18 s without interaction on screen. The map could also be closed via pressing an “ok” button. After closing the map, the walking phase was initiated, where only footsteps were shown on screen together with two response buttons: a “I arrived” button, which initiated the capture phase when pressed, and a “I dońt know where I am” button, which initiated the help phase. When arriving at the POI, the participant had to press the “I arrived” button, which opened the phone camera, to scan a QR-Code that was placed at the entrance door of the building (capture phase). The “I don’t know where I am” button could be pressed when the participant felt lost and wanted to see the map again (help phase). After successfully scanning the QR code, the same procedure was initiated for the next POI. GPS data (latitude, longitude, and timestamp) were recorded by the app every two seconds, together with certain meta-data, such as the number of map views during walking. During the start and help phases as well as during the capture phases, sensor data (gyroscope and accelerometer) were recorded in addition, which are not considered here. The data were saved encrypted on the phone and were decrypted after downloading with the customized “Explore App decryption tool”, also provided by Intenta GmbH.

Before testing, participants performed a short practice track where they had to walk from a nearby parking lot to the DZNE. For this track, they were guided by the experimenter who introduced the task procedure and practiced all app features with them. Afterwards, participants were instructed to find the five POIs independently, while using the help function of the app only when absolutely necessary. After returning to the DZNE, participants were debriefed and asked if they used certain strategies during task performance.

### Data preprocessing

All data were analyzed using R v4.1.1 (R Core Team, 2021). The GPS data were first cleaned by removing duplicated entries and checked for data artifacts, for example, when the GPS signal was lost. Data for nine tracks from eight participants (1 younger adult, 5 healthy older adults, 2 older adults with SCD) were removed due to technical problems or difficulties in following task instructions. For example, one participant reported in the debriefing, that she asked other pedestrians for directions on some tracks and the data for these tracks showed that she stopped at popular meeting spots. Hence, these tracks were excluded from the analyses. Three older adults with SCD discontinued participation prior to completing the round for various reasons (e.g., weather change or tiredness), resulting in incomplete data with five missing tracks for them. We further removed data that were recorded 15 m around the location of the POIs, when the QR code sign could already be seen. Data from the last track, where participants returned to the DZNE from the last POI, were not considered in the analyses because the DZNE also served as the starting point of the task. The five tracks in our analysis varied in length and number of decision points (intersections at which the participants could choose between different directions). The minimum distance that had to be covered for track 1 was approximately 147 m with five decision points (when following the most direct path). Track 2 covered 67 m with three decision points. Track 3 was the longest with 258 m minimum path length and nine decision points. Track 4 covered 149 m with four decision points and track 5 202 m with six decision points. Overall, 346 tracks were included in the analyses that considered data from the whole sample. For some of the analyses (GPS data analysis, LPA), only data from those participants were considered who completed all five tracks of the task (23 younger adults; 20 healthy older adults; 18 older adults with SCD; see Supplementary Table 7 for the characteristics of the sub-sample), resulting in 305 tracks that were analyzed. The sub-sample showed similar characteristics as the whole sample in terms of age, gender distribution, and our questionnaire measures (e.g., the groups did not differ with respect to campus familiarity and general mobility, all p ≥ .272).

### GPS data analysis

To quantify differences in the movement trajectories between participants, we applied dynamic time warping (DTW) to the GPS data that were recorded during the walking phases, using the *dtw* package (Giorgino, 2009). We only considered data from those participants who completed all tracks of the task because otherwise participants who completed the same subset of tracks would be considered more similar relative to those who completed all tracks. DTW has been used in previous studies examining movement trajectories on a similar time scale (Haase & Brefeld, 2013), it is suitable for time-series that vary in length, and proved to be robust to outliers (Cleasby et al., 2019; Senin, 2009). The resulting DTW dissimilarity matrix was normalized using a Min-Max normalization (range: 0-1; Patro & Sahu, 2015) and then served as input feature in a k-medoid clustering analysis, as implemented in the *cluster* package (Maechler et al., 2012). We varied the number of possible clusters from three to seven and computed the average Silhouette coefficient as a measure for the distance between the resulting clusters (possible range: –1 to 1 with negative values indicating wrong cluster assignments and values near zero overlapping clusters). We found three clusters to be the best choice for our sample (Silhouette coefficients per tested cluster number: 3 = 0.48; 4 = 0.28; 5 = 0.27; 6 = 0.31; 7 = 0.32). Next, the correspondence between the wayfinding clusters and participant classes was determined by calculating the cluster purity (possible range: 0-1 with higher values indicating lower within-cluster variation in terms of class labels).

### Performance measures analysis

For each track and participant, we extracted five performance measures from the GPS and user input data. First, the wayfinding distance was calculated by summarizing the haversine distance of adjacent coordinates. Second, the wayfinding duration was calculated as the sum of temporal differences between adjacent coordinates. Third, the movement speed was calculated by dividing the wayfinding distance by the wayfinding duration. Fourth, the number of help function calls during walking (map views) was extracted from the meta-data. Lastly, we calculated the number of orientation stops, defined as the number of times when the participants moved less than one meter in five seconds during walking, using the stay point algorithm (Montoliu et al., 2013). Orientation stops have been shown to be suggestive of additional cognitive processing demands during navigation, for example, when participants have difficulties in finding a goal and need to retrieve relevant spatial information in order to solve the task (May et al., 1995; Taillade et al., 2013).

To identify subgroups of navigators in our sample based on these performance measures, we performed a latent profile analysis (LPA) with the z-transformed scores of each measure, averaged across tracks, as input parameters (Berlin et al., 2014). Again, we only included data from those participants who completed all tracks of the task to allow a better comparability to the results of the GPS data analysis and because we used the mean scores across tracks for the analysis. LPA is also a data-driven approach that aims at the extraction of homogenous sub-samples and the resulting wayfinding profiles can be examined with respect to their performance characteristics. We fitted models with three to seven profiles to the data and evaluated them by comparing the Akaike information criterion (AIC), the Bayesian information criterion (BIC), the sample-adjusted BIC (SABIC), the bootstrap likelihood ratio test (BLRT), entropy values, and the number of observations in each profile (see Table 2; Berlin et al., 2014). The AIC was lowest for the model with seven profiles, whereas the BIC was lowest for the three-profile model. The lowest SABIC value was found for the seven-profile model. Entropy, which can vary between zero and one with higher values corresponding to better classification, was above 0.9 for models with four and more profiles. The bootstrap likelihood ratio test was not significant for the models with four, five, and six profiles. Hence, those models were excluded from further consideration. For the seven-profile model, more than three profiles contained 6 or fewer members, which were not deemed reliable (Lubke & Neale, 2006). Thus, the model with three wayfinding performance profiles was accepted. Again, we calculated the cluster purity to determine how well these profiles represented our participant classes.

**Table 2.**
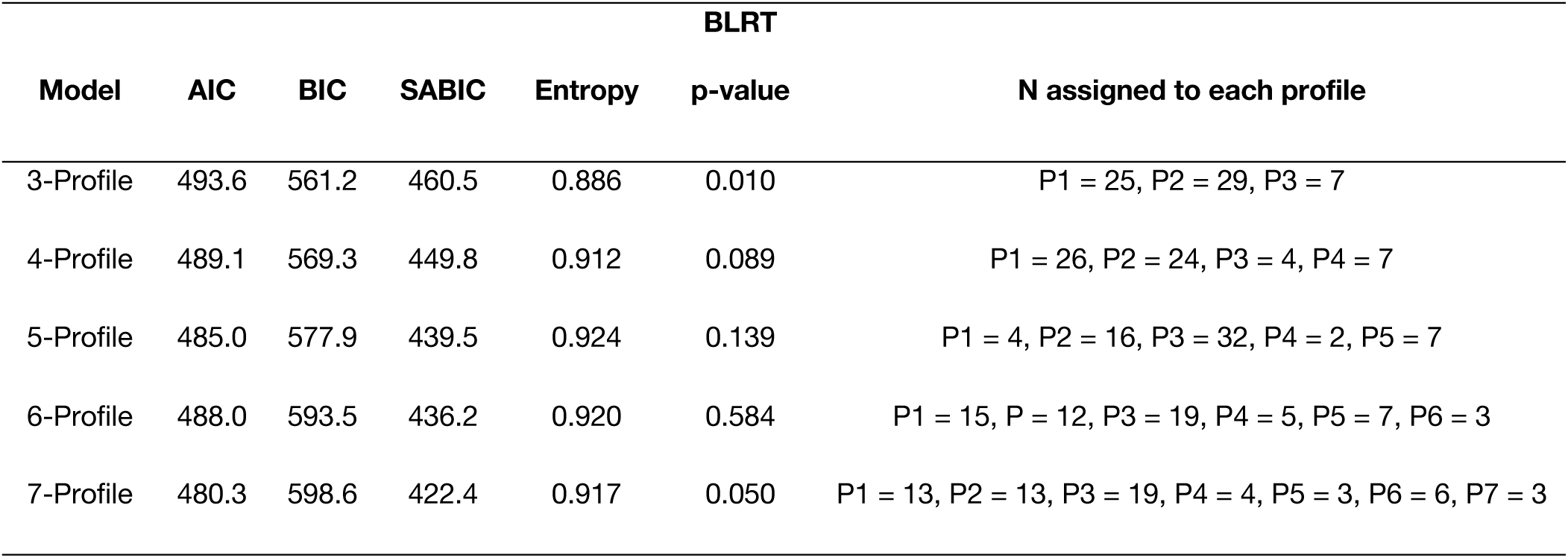
Model fit statistics of the latent profile analysis (LPA) with five performance measures as input features (wayfinding distance; wayfinding duration; movement speed; number of map views; number of orientation stops) for models with three to seven profiles. AIC = Akaike information criterion; BIC = Bayesian information criterion; SABIC = sample-adjusted Bayesian information criterion; BLRT = bootstrap likelihood ratio test.

We then fitted, for each performance measure, mixed effect models to the data, allowing us to assess group differences in more detail. For this analysis, data from the whole sample were considered. For the log-transformed wayfinding distance and duration as well as the movement speed, we fitted a LME model to the data, using the *lme4* package (Bates et al., 2014). For the number of map views during wayfinding and the number of orientation stops, GME models were fitted to the data, as implemented in the *glmmTMB* package (Brooks et al., 2017) due to zero-inflation of the data. All models were estimated with gender and campus familiarity as covariates and track and participant as random intercepts (basic model). The fixed effect group (younger adults, healthy older adults, and older adults with SCD) was then added to estimate the final model. The two models were compared by using the AIC with lower AIC values indicating better model fit (Burnham & Anderson, 2004; Meteyard & Davies, 2020). The significance of all fixed effect predictors in the LME models was assessed using t-tests and the Satterthwaite’s approximation for degrees of freedom. The significance of the fixed effect predictors in the GME models was tested using z-tests. Detailed information on the results of the mixed effect models is provided in Supplementary Tables 1-5.

Next, we used the average number of orientation stops across tracks, which was the performance measure on which older adults with SCD differed from healthy older adults, as predictor in a logistic regression model with SCD status as outcome variable in the subsample of older participants (n = 48). The prediction accuracy for data from unknown participants was assessed in a LOO cross-validation by fitting a logistic regression model in all but one participant and evaluating the prediction of the model parameter in the left-out (test) participant. This process was repeated 48 times so that each participant served as a test participant once. Afterwards, the diagnostic accuracy was assessed in a receiver operating characteristics (ROC) analysis with the *pROC* package (Robin et al., 2011). The optimal cut-off values were determined by either maximizing sensitivity or specificity while constraining the other metric to a minimum of 0.5, using the *cutpointr* package (Thiele & Hirschfeld, 2020).

As a last step, we calculated Pearson product-moment correlation coefficients between the number of orientation stops and the age-, gender-, and education-corrected z-scores from all available subtests of the CERAD test battery (Welsh et al., 1994) for the older adults with SCD (n= 23) to provide indications about which cognitive processes might be associated with this performance measure.

### Data availability

The corresponding author (JM) will provide the source data files upon reasonable request.

### Code availability

The corresponding author (JM) will provide the code for the analysis of the source data upon reasonable request.

## Data Availability

All data produced in the present study are available upon reasonable request to the authors.

## Acknowledgements

This work was funded by the Deutsche Forschungsgemeinschaft (DFG, German Research Foundation) – Project-ID 425899996 – SFB 1436 and a DZNE Innovation 2 Application Award (awarded to Nadine Diersch; 2019-2021).

We thank Jan Schlosshauer (Head of R&D) and his development team at Intenta GmbH for the great cooperation and the realization of the “Explore” app. We further thank Vincent Koepp for supporting the initial app development stages and his assistance during the development and piloting of the familiarity questionnaire, Celine Winter and Annemarie Scholz for their assistance with data collection, and Vishnu Unnikrishnan for his assistance with the data analyses.

## Author contributions

Conceptualization, N.D.; Project administration, J.M. and N.D.; Investigation, J.M.; Resources, M.B., W.G., and N.D.; Methodology & Formal Analysis, J.M., P.M., and M.S.; Visualization, J.M. and N.D.; Writing – Original Draft, J.M.; Writing – Review & Editing, S.S., E.K., A.M., and N.D.; Supervision, A.M. and N.D.; Funding Acquisition, S.S., E.K., A.M., and N.D.

