## Supplementary Information for "Identifying older adults at risk for Alzheimer’s Disease based on smartphone data obtained during wayfinding in the real world"

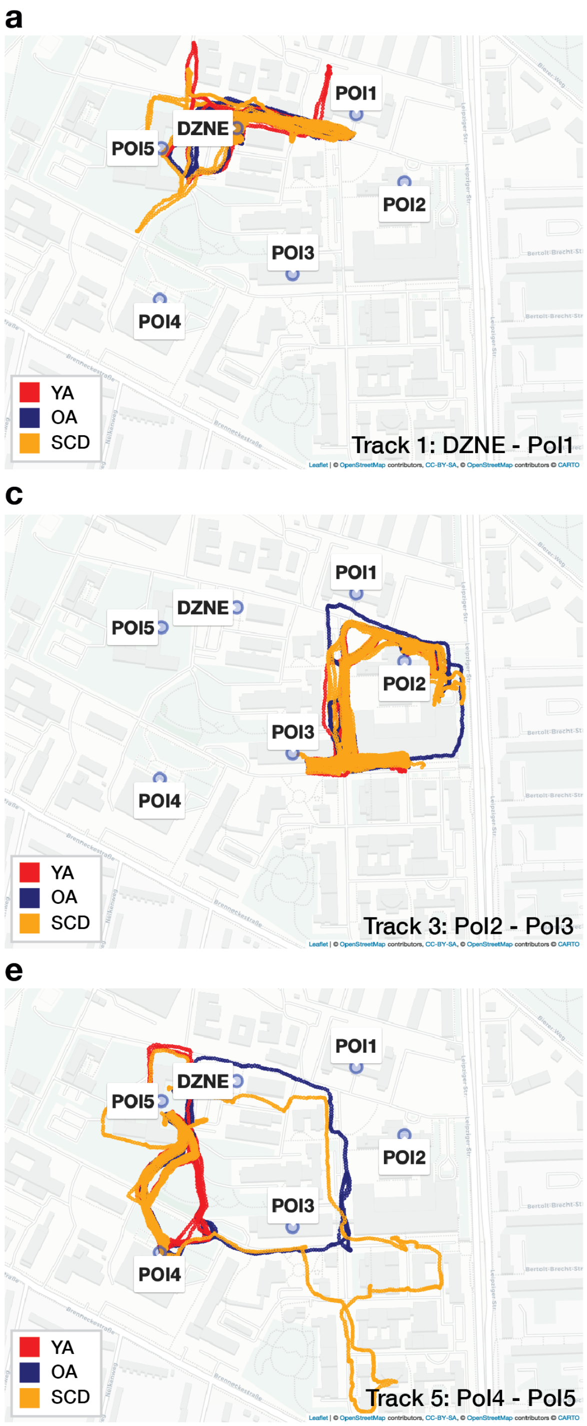


**Supplementary Figure 1.** Movement trajectories of the three participant groups (red: younger adults; blue: healthy older adults; yellow: older adults with subjective cognitive decline) on (a) track 1; (b) track 2; (c) track 3; (d) track 4; (e) track 5 of the mobile wayfinding task.

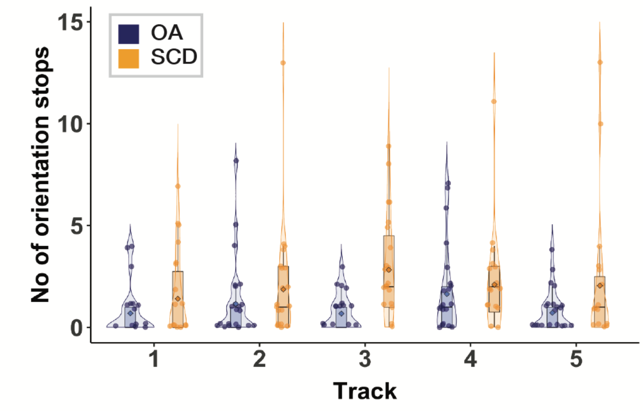


**Supplementary Figure 2.** Number of orientation stops on each track in healthy older adults (blue) and older adults with subjective cognitive decline (yellow). The boxplot denotes the lower and upper quartile of the measure; center line the median; whiskers the 1.5x interquartile range; dots the individual data points; diamond shape the mean.

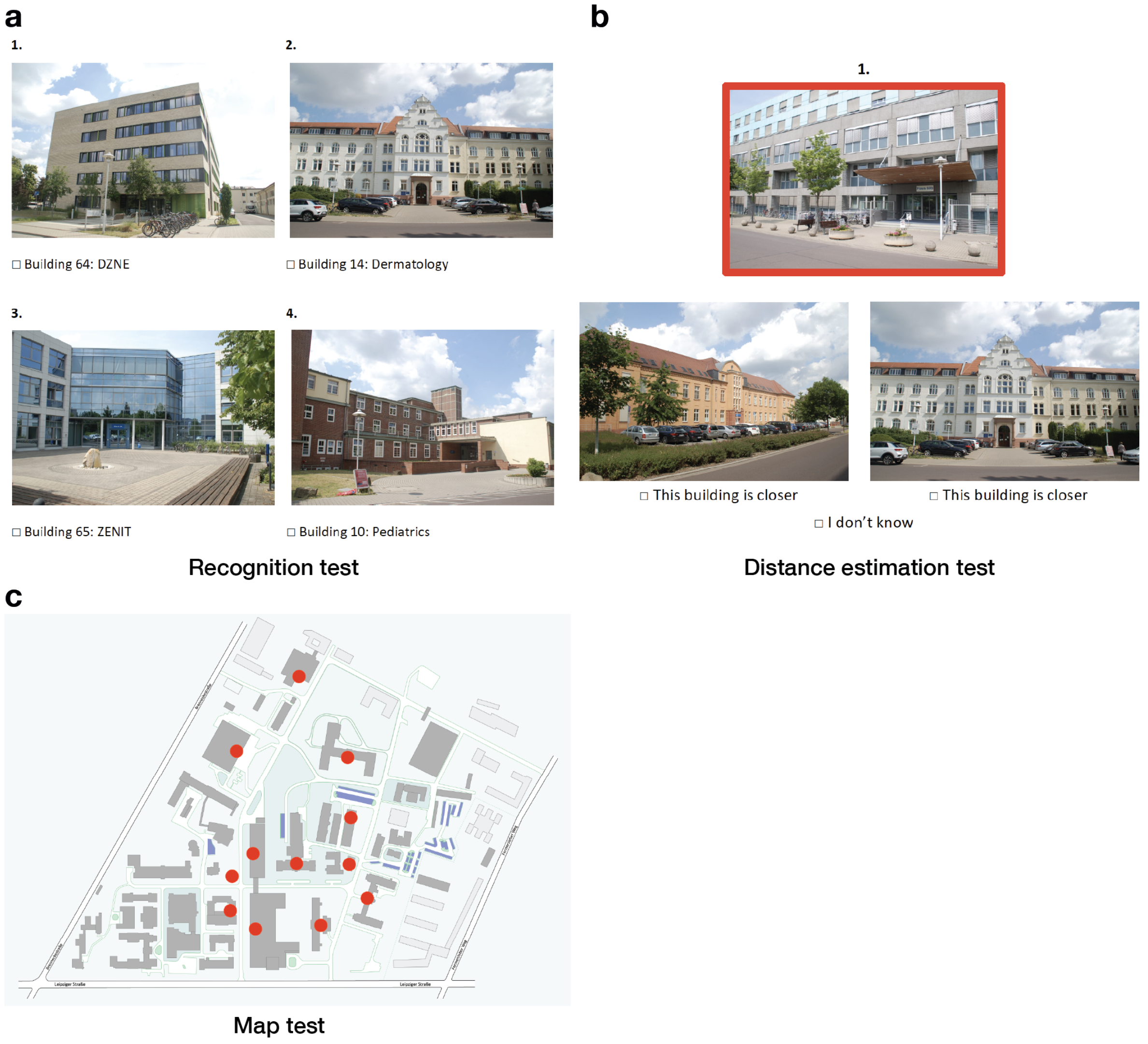


**Supplementary Figure 3.** Spatial memory test implemented in the familiarity questionnaire to assess the participants' prior knowledge of the campus area in three sub-tests (maximum score: 28). (a) Recognition test: First, participants had to indicate from a list of pictures showing 12 campus buildings (including the 5 PoIs of the mobile wayfinding task), which of the buildings they recognize (shown are 4 example buildings). (b) Distance estimation test: Next, they saw 4 triplets of the 12 buildings and were asked to indicate, which of the two buildings in the lower row lies closer to the reference building in the upper row (shown is one example triplet). (c) Map test: Finally, for the buildings they knew, they had to assign the buildings from the recognition test to dots on a map of the campus, in this way identifying their location.

| | | | | | |
|---|---|---|---|---|---|
| | | **Fixed Effects** | | | |
| | Est/Beta | SE | 95% CI | t-value | p |
| **Intercept** | 5.368 | 0.184 | 5.007 ; 5.729 | 29.136 | **< .001** |
| **Group YA** | -0.127 | 0.052 | -0.230 ; -0.024 | -2.427 | **.018** |
| **Group SCD** | 0.071 | 0.055 | -0.038 ; 0.179 | 1.273 | .208 |
| **Familiarity** | -0.003 | 0.003 | -0.009 ; 0.002 | -1.241 | .219 |
| **Gender Female** | 0.102 | 0.002 | 0.016 ; 0.188 | 2.333 | **.023** |
| | | **Random Effects** | | | |
| | | Variance | SD | | |
| **Participant** | | 0.003 | 0.055 | | |
| **Track** | | 0.154 | 0.393 | | |
| | | **Model fit** | | | |
| **AIC** | | Basic Model | Final Model | | Delta AIC |
| | | 370.34 | 361.09 | | -9.25 |
| **$R^2$** | | | Marginal | | Conditional |
| | | | 0.033 | | 0.530 |

**Supplementary Table 1.** Results of the linear mixed effect model estimating the fixed effects of group, campus familiarity, and gender on the log-transformed wayfinding distance. Random intercepts were estimated per participant and track. P-values for fixed effects were calculated using the Satterthwaite's approximation for degrees of freedom (bold font indicates significant effects). Confidence intervals were calculated using the Wald method. Model equation: log(wayfinding distance) ~ group + familiarity + gender + (1|participant) + (1|track).

| Fixed Effects | | | | | |
|---|---|---|---|---|---|
| | Est/Beta | SE | 95% CI | t-value | p |
| **Intercept** | 5.120 | 0.178 | 4.771 ; 5.468 | 28.802 | **< .001** |
| **Group YA** | -0.283 | 0.070 | -0.420 ; -0.146 | -4.042 | **< .001** |
| **Group SCD** | 0.128 | 0.074 | -0.017 ; 0.273 | 1.733 | *.088* |
| **Familiarity** | -0.008 | 0.004 | -0.015 ; -0.001 | -2.114 | **.038** |
| **Gender Female** | 0.139 | 0.058 | 0.025 ; 0.253 | 2.391 | **.020** |
| **Random Effects** | | | | | |
| | | Variance | SD | | |
| **Participant** | | 0.022 | 0.148 | | |
| **Track** | | 0.130 | 0.361 | | |
| **Model fit** | | | | | |
| **AIC** | | Basic Model | Final Model | | Delta AIC |
| | | 483.35 | 458.01 | | -25.34 |
| **R²** | | | Marginal | | Conditional |
| | | | 0.108 | | 0.512 |

**Supplementary Table 2.** Results of the linear mixed effect model estimating the fixed effects of group, campus familiarity, and gender on the log-transformed wayfinding duration. Random intercepts were estimated per participant and track. P-values for fixed effects were calculated using the Satterthwaite's approximation for degrees of freedom (bold font indicates significant effects; italic font indicates statistical trends). Confidence intervals were calculated using the Wald method. Model equation: log(wayfinding duration) ~ group + familiarity + gender + (1|participant) + (1|track).

| Fixed Effects | | | | | |
|---|---|---|---|---|---|
| | Est/Beta | SE | 95% CI | t-value | p |
| **Intercept** | 1.310 | 0.058 | 1.196 ; 1.424 | 22.504 | **< .001** |
| **Group YA** | 0.223 | 0.051 | 0.123 ; 0.323 | 4.375 | **< .001** |
| **Group SCD** | -0.048 | 0.053 | -0.153 ; 0.056 | -0.901 | .371 |
| **Familiarity** | 0.006 | 0.003 | 0.001 ; 0.011 | 2.206 | **.031** |
| **Gender Female** | -0.054 | 0.042 | -0.137 ; 0.029 | -1.285 | .203 |
| **Random Effects** | | | | | |
| | | Variance | SD | | |
| **Participant** | | 0.028 | 0.168 | | |
| **Track** | | 0.002 | 0.130 | | |
| **Model fit** | | | | | |
| **AIC** | | Basic Model | Final Model | | Delta AIC |
| | | -229.02 | -252.34 | | -23.32 |
| **$R^2$** | | | Marginal | | Conditional |
| | | | 0.277 | | 0.742 |

**Supplementary Table 3.** Results of the linear mixed effect model estimating the fixed effects of group, campus familiarity, and gender on the movement speed. Random intercepts were estimated per participant and track. P-values for fixed effects were calculated using the Satterthwaite's approximation for degrees of freedom (bold font indicates significant effects). Confidence intervals were calculated using the Wald method. Model equation: movement speed ~ group + familiarity + gender + (1|participant) + (1|track).

| Fixed Effects | | | | | |
|---|---|---|---|---|---|
| | Est/Beta | SE | 95% CI | z-value | p |
| **Intercept** | -0.655 | 0.506 | -1.646 ; 0.336 | -1.296 | .195 |
| **Group YA** | -1.243 | 0.514 | -2.251 ; -0.236 | -2.418 | **.016** |
| **Group SCD** | 0.820 | 0.435 | -0.033 ; 1.672 | 1.885 | *.059* |
| **Familiarity** | -0.048 | 0.025 | -0.010 ; -0.000 | -1.960 | **.050** |
| **Gender Female** | 0.183 | 0.367 | -0.537 ; 0.903 | 0.499 | .618 |
| **Random Effects** | | | | | |
| | | Variance | SD | | |
| **Participant** | | 1.246 | 1.116 | | |
| **Track** | | 0.003 | 0.063 | | |
| **Model fit** | | | | | |
| **AIC** | | Basic Model | Final Model | | Delta AIC |
| | | 661.5 | 647.4 | | -14.1 |
| **Pseudo $R^2$** | | | 0.265 | | |

**Supplementary Table 4.** Results of the generalized mixed effect model estimating the fixed effects of group, campus familiarity, and gender on the number of map views during walking. Random intercepts were estimated per participant and track. The model was estimated using the maximum likelihood estimation (bold font indicates significant effects; italic font indicates statistical trends). A zero-inflated negative binomial distribution of the data was assessed and a log link function applied. Confidence intervals were calculated using the Wald method. Pseudo $R^2$ was calculated by: 1-e^((-2/n*logL(x)-logL(0))), where logL(x) is the is the log-likelihood of the final model and logL(0) the log-likelihood of an intercept only model. Model equation: number of map views ~ group + familiarity + gender + (1|participant) + (1|track).

| | Est/Beta | SE | 95% CI | z-value | p |
|---|---|---|---|---|---|
| **Fixed Effects** | | | | | |
| **Intercept** | 0.187 | 0.291 | -0.384 ; 0.758 | 0.641 | .521 |
| **Group YA** | -0.919 | 0.281 | -1.470 ; -0.369 | -3.274 | **.001** |
| **Group SCD** | 0.670 | 0.248 | 0.184 ; 1.157 | 2.702 | **.006** |
| **Familiarity** | -0.007 | 0.013 | -0.033 ; 0.019 | -0.531 | .595 |
| **Gender Female** | 0.301 | 0.207 | -0.105 ; 0.707 | 1.453 | .146 |
| **Random Effects** | | | | | |
| | | Variance | SD | | |
| **Participant** | | 0.402 | 0.634 | | |
| **Track** | | 0.000 | 0.000 | | |
| **Model fit** | | | | | |
| **AIC** | | Basic Model | Final Model | | Delta AIC |
| | | 1020.1 | 994.1 | | -25.94 |
| **Pseudo $R^2$** | | | 0.268 | | |

**Supplementary Table 5.** Results of the generalized mixed effect model estimating the fixed effects of group, campus familiarity, and gender on the number of orientation stops. Random intercepts were estimated per participant and track. The model was estimated using the maximum likelihood estimation (bold font indicates significant effects). A zero-inflated poisson distribution of the data was assessed and a log link function applied. Confidence intervals were calculated using the Wald method. Pseudo $R^2$ was calculated by: 1-e^((-2/n*logL(x)-logL(0))), where logL(x) is the is the log-likelihood of the final model and logL(0) the log-likelihood of an intercept only model. Model equation: number of orientation stops ~ group + familiarity + gender + (1|participant) + (1|track).

| CERAD Subtest | r | p-value |
|---|---|---|
| Verbal Fluency (animals) | -.10 | .640 |
| Boston Naming Test | .20 | .371 |
| Mini-Mental Status Examination | -.33 | .122 |
| Word List Learning | -.26 | .228 |
| Word List Recall | -.18 | .412 |
| Word List Savings | .03 | .901 |
| Word List Discrimination | .14 | .524 |
| Constructional Praxis Drawing | .21 | .332 |
| Constructional Praxis Recall | -.32 | .136 |
| Constructional Praxis Savings | -.31 | .144 |
| Verbal Fluency (words)[a] | .16 | .494 |
| Trail Making Test A | -.07 | .756 |
| Trail Making Test B | -.29 | .184 |
| Trail Making Test A/B | -.39 | .068 |

**Supplementary Table 6.** Pearson product-moment correlation coefficients (df = 21) between the number of orientation stops and the age-, gender-, and education-corrected z-scores from all available subtests of the CERAD test battery in older adults with subjective cognitive decline (SCD). [a] three missing values for Verbal Fluency (words), df = 18.

| | YA | OA | SCD |
|---|---|---|---|
| n | 23 | 20 | 18 |
| Age | 24.4 ± 2.29 | 66.0 ± 3.79 | 65.2 ± 6.87 |
| No of female | 12 | 9 | 10 |
| Campus familiarity (max. score: 28) | 12.9 ± 9.30 | 13.4 ± 8.40 | 9.2 ± 6.59 |
| Life-Space Assessment (max. score: 120) | 81.3 ± 12.3 | 84.8 ± 16.1 | 83.4 ± 14.1 |
| Cognitive screening scores | -- | MoCA: 28.4 ± 1.09 | MMSE: 29.0 ± 1.19<br>CERAD: 0.24 ± 0.54<br>min = -0.75 |

**Supplementary Table 7.** Sample characteristics (descriptives, mean scores ± SD) of the three participant groups (YA: younger adults; OA: healthy older adults; SCD: older adults with subjective cognitive decline), when only considering those individuals who completed all five tracks in the mobile wayfinding task (N = 61). The CERAD composite score was calculated using the age-, gender-, and education-corrected z-scores from six different subtests (Boston Naming Test, verbal fluency, word list learning, word list recall, word list savings, and constructional praxis, see Chandler et al., 2005). The groups did not differ in the listed attributes, all $p \geq .272$ (age differences were only tested between healthy older adults and older adults with SCD).
